# Trans-ethnic Genomic Informed Risk Assessment for Alzheimer’s disease: An International Hundred K+ Cohorts Consortium Study

**DOI:** 10.1101/2022.12.22.22283738

**Authors:** Patrick M. Sleiman, Hui-Qi Qu, John J Connolly, Frank Mentch, Alexandre Pereira, Paulo A Lotufo, Stephen Tollman, Ananyo Choudhury, Michele Ramsay, Norihiro Kato, Kouichi Ozaki, Risa Mitsumori, Jae-Pil Jeon, Chang Hyung Hong, Sang Joon Son, Hyun Woong Roh, Dong-gi Lee, Naaheed Mukadam, Isabelle F Foote, Charles R Marshall, Adam Butterworth, Bram P Prins, Joseph T Glessner, Hakon Hakonarson, the Davos Alzheimer Collaborative and IHCC consortium

**Affiliations:** The Center for Applied Genomics, Children’s Hospital of Philadelphia, Philadelphia, Pennsylvania, 19104, USA; Department of Pediatrics, The Perelman School of Medicine, University of Pennsylvania, Philadelphia, Pennsylvania, 19104, USA; Division of Human Genetics, Children’s Hospital of Philadelphia, Philadelphia, Pennsylvania, 19104, USA; Faculdade de Medicina da Universidade de São Paulo, São Paulo, Brazil; Centro de Pesquisas Clínicas e Epidemiológicas, Hospital Universitário, Universidade de São Paulo, São Paulo, Brazil; MRC/Wits Rural Public Health and Health Transitions Research Unit (Agincourt), School of Public Health, Faculty of Health Sciences, University of the Witwatersrand, Johannesburg, South Africa; Sydney Brenner Institute for Molecular Bioscience, Faculty of Health Sciences, University of the Witwatersrand, Johannesburg, South Africa; National Center for Global Health and Medicine, Tokyo, 1628655, Japan; Medical Genome Center, Research Institute, National Center for Geriatrics and Gerontology (NCGG), Obu City, Aichi Prefecture, Japan; Korea Biobank Project, Korea National Institute of Health, Osong, Korea; Department of Psychiatry, Ajou University School of Medicine, Suwon, Korea; Preventive Neurology Unit, Wolfson Institute of Population Health, Queen Mary University of London, UK; Genes & Health, Blizard Institute, Queen Mary University of London, UK; MRC/BHF Cardiovascular Epidemiology Unit, Department of Public Health and Primary Care, University of Cambridge, Cambridge, UK; NIHR Blood and Transplant Research Unit in Donor Health and Genomics, Department of Public Health and Primary Care, University of Cambridge, Cambridge, UK; Division of Pulmonary Medicine, Children’s Hospital of Philadelphia, Philadelphia, Pennsylvania, 19104, USA; Faculty of Medicine, University of Iceland, 101 Reykjavik, Iceland

**Keywords:** Alzheimer’s disease, data sharing, female infertility, genomic informed risk assessment, polygenic risk score, minority population, thyroid, trans-ethnic

## Abstract

**Background:** Alzheimer’s disease (AD) is a complex multifactorial progressive dementia affecting all human populations. As a collaboration model between the International Hundred K+ Cohorts Consortium (IHCC) and the Davos Alzheimer Collaborative (DAC), our aim was to develop a trans-ethnic genomic informed risk assessment (GIRA) algorithm for AD.

**Methods:** The GIRA model was created to include a polygenic risk score (PRS) calculated from the AD GWAS loci, the *APOE* haplotypes, and non-genetic covariates including age, sex and first 3 principal components of population substructure. The model was first validated using a ancestrally diverse dataset from the eMERGE network, and subsequently validated in a South-Asian population in the UK and 3 East-Asian populations. The distributions of the PRS scores were also explored in populations from 3 African regions. In two validation sites, the PRS was tested for associated with the levels of plasma proteomics markers.

**Results:** We created a trans-ethnic GIRA model for the risk prediction of AD and validated the performance of the GIRA model in different populations. The proteomic study in the participant sites identified proteins related to female infertility and autoimmune thyroiditis and associated with the risk scores of AD, highlighting molecular mechanisms underlying the previously observed correlations between these clinical phenotypes.

**Conclusions:** As the initial effort by the IHCC to leverage existing large scale datasets in a collaborative setting with DAC, we developed a trans-ethnic GIRA for AD with the potential of identifying individuals at high risk of developing AD for future clinical applications. The PRS scores in this model also contribute new research discoveries for the molecular pathogenesis of AD as demonstrated by the proteomic data.

## Introduction

Alzheimer’s disease (AD) is characterized by irreversible neuronal degeneration with no cure to date^1^. AD affects all human populations with the prevalence of 8.4% to 13.8% in people aged 65 years and older according to the Centers for Disease Control and Prevention (CDC, https://www.cdc.gov/media/releases/2018/p0920-alzheimers-burden-double-2060.html). Early diagnosis of AD may enable early intervention to minimize the damage to the central nervous system^2^. With older age and female sex as the known risk factors^3^, AD has been recognized as a complex multifactorial genetic disease^4^. The apolipoprotein E gene (*APOE*) variants have been established as a major susceptibility locus for AD^5^. APOE is a major apoprotein of the lipoproteins chylomicron and very low density lipoproteins (VLDL), with the function of lipid transportation^6^. Amyloid β (Aβ) forms the extracellular plaque in AD brains, and is the central mechanism of the AD pathogenesis^7^. The APOE is associated with Aβ aggregation and clearance by physical interaction and sharing common receptors^8^.

Human *APOE* has three common genetic isoforms APOE2 (112Cys-158Cys), APOE3 (112Cys-158Arg), and APOE4 (112Arg-158Arg), with different amino acid residues at positions 112 and 158 in the N-terminal domain^8 9^. The APOE genetic isoforms are determined by two single nucleotide polymorphisms: (1) rs429358 (hg38, chr19:44908684:T:C) causes amino acid substitution of Cys to Arg at residue 112; (2) rs7412 (hg38, chr19:44908822:C:T) causes amino acid substitution of Arg to Cys at residue 158^10^. Among the 3 APOE isoforms encoded by 3 haplotypes of rs429358 and rs7412 SNPs, the APOE3 haplotype has the most common frequency in all populations studied to date, including African populations ^11 12^. Compared to the APOE3 haplotype, APOE2 is protective against AD, and APOE4 is predisposing to AD with an odds ratio (OR) value about 2 to 4 for heterozygous carriers^11 13 14^. Despite the large genetic effect, however, the *APOE* locus alone is insufficient to explain the genetic susceptibility to AD. A large number of genetic loci associated with AD, e.g. WASH complex subunit 3 (*WASHC3*), bridging integrator 1 (*BIN1*), complement C3b/C4b receptor 1 (Knops blood group) (*CR1*), and cathepsin B (*CTSB*), have recently been identified in the genome-wide association studies (GWAS) by Jun et al.^15^ and Bellenguez et al.^16^ with large sample sizes including participants with different ancestries. Additional genetic information from these AD loci may improve the risk prediction of AD and the identification of patients with high risk of AD.

In this study, we developed a genomic informed risk assessment (GIRA) algorithm for AD with both the *APOE* isoforms and the polygenic risk score (PRS) calculated by the multiple AD loci identified in previous GWAS^15 16^, as well as known non-genetic risk factors of AD including age and sex. The PRS aggregated the effects of GWAS loci other than the *APOE* locus into a single score, an approach that has been shown as an effective approach to identify patients with high risk of many complex diseases^17-19^. In particular, a PRS with selected loci has been demonstrated for precise prediction of type 1 diabetes (T1D)^20^. Compared to AD with *APOE* as a major genetic locus plus a number of minor loci, T1D has also the human leukocyte antigen (*HLA*) region as a major genetic locus plus a number of minor loci^20^. Enabled by risk prediction using GIRA, and with more attention given to the high risk patients, this will consequently enable early diagnosis and early intervention for the disease. Nowadays, the major challenge in GIRA models is potential health disparities due to the lack of genomic information in minority or understudied populations^21^. In many cases, effective GIRA models relying on data from large-sample genome-wide association studies (GWAS) are only available for the European population as the majority of GWASs have been done in people with European ancestry^22^. With the aim to eliminate genomics-related health disparities, we leveraged an international effort supported by the International HundredK+ Cohorts Consortium (IHCC)^23^, which has brought together large scale cohorts of diverse populations from around the world, who have genome-wide genotype data.

## Methods

### Genotyping

The genotyping data was imputed with the TOPMed (Version R2 on GRC38) Reference Panel at the TOPMed imputation server (https://imputation.biodatacatalyst.nhlbi.nih.gov), or the Haplotype Reference Consortium (HRC) panel at the Michigan imputation server (https://imputationserver.sph.umich.edu), or the Sanger Imputation Server (African reference panel), at each participant site. No quality issue was reported for the imputation of any of the SNP markers in the GIRA model from any of participating sites. In case of quality issues for the imputation, to find alternative SNP markers based on the specific population and the genotyping arrays would be done in house upon request.

### *APOE* haplotyping

The *APOE* haplotypes were inferred from the two SNPs rs429358 and rs7412, i.e. APOE2=rs429358/T-rs7412/T; APOE3=rs429358/T-rs7412/C; APOE4=rs429358/C-rs7412/C.

### GWAS loci

The GWAS loci were identified by the Stage I studies from Jun et al.^15^ and Bellenguez et al.^16^ The study by Jun et al. included cases/controls with European ancestry (EA, n=13,100/13,220), African Americans (AA, n=1,472/3,511), Japanese (JPN, n=951/894), and Israeli-Arab (IA, n=51/64)^15^. The study by Bellenguez et al. included 2,447 diagnosed cases, 46,828 proxy cases of dementia and 338,440 controls, all with European ancestry^16^. Multi-allelic variants, indels and rare SNPs with MAF < 3% were excluded from further analysis. The remaining variants from the combined summary stats were LD pruned using an R^2^ threshold of 0.3 resulting in a final list of 74 variants (Supplementary Table 1).

### GIRA model

The GIRA model includes 3 components, i.e. *APOE* haplotype, PRS by genomic markers, and covariates including age, sex and the first 3 principal components of population substructure for genetic ancestry correction. The PRS is calculated by the weighted sum of the effect alleles, while weights are the reported odds ratios.

### Validation of PRS models

The PRS markers were first validated for their allele frequencies in 4145 ancestrally diverse dementia patients from the electronic medical records and genomics (eMERGE) consortium Phase I-III dataset. Subsequently, the scoring algorithm, SNPs and weights, as well as detailed instructions were disseminated to participant sites (Table 1). Scores were generated on a total of 25,786 participants across a variety of endpoints and returned to the Children’s Hospital of Philadelphia (CHOP) for collation. Each site was requested to follow standardized reporting metrics. As all groups did not have accurate age at onset data, we requested odds ratios rather than hazard ratios for the phenotype outcomes (Table 1). Each validation site was required to return:

**Table 1.** Sites participating in the validation.

| Site | Genetic ancestry | Phenotypic outcome | N (case/control) |
| --- | --- | --- | --- |
| National Center for Geriatrics and Gerontology (NCGG) Biobank | Japanese (East Asian) | AD, MCI | Case:1000 Normal Cognitive:1000 |
| East London Genes and Health cohort | British-Pakistani/British-Bangladeshi (South Asian) | All-cause dementia (from secondary/primary care records); MCI/cognitive decline cases excluded | 104 cases; 614 controls |
| Korean Biobank Project | Korean (East Asian) | Phenotype 1: Cortical amyloid positivity (by Flutemetamol PET imaging) (Control: Cortical amyloid negativity) | 191:337 (total 528) |
|  |  | Phenotype 2: CDR global Score 1 or over (Control: CDR global 0.5 or less) | 157:539 (total 696) |
| Africa Wits-INDEPTH partnership for genomic studies (AWI-Gen) | African (Different ethnolinguistic and geographic groups) | Not assessed for AD | Population cross sectional cohort: n=10,700 |
| The Brazilian Longitudinal Study of Adult Health (ELSA-Brasil) | Brazilian (Admixed) |  |  |
| INTERVAL (UK Blood Donors) | European (White British) | Stroop Test (attention and reaction times), Trail Making Test (executive function), Pairs Test (Episodic Memory), Reasoning Tests (intelligence), >3K proteins on the SomaLogic proteomics platform | ~9k Cognitive measure; 1140 proteomics |
| UKBiobank | European (White British) | ICD9/10 diagnoses of AD/dementia derived from hospital inpatient records or patients with date of Alzheimer's disease/dementia report | 2923 cases; 226342 controls |

Abbreviations: AD, Alzheimer's disease; CDR, Clinical Dementia Rating; ICD, International Classification of Diseases; MCI, mild cognitive impairment; NA, not applicable.

1. The odds ratio per standard deviation of the PRS distribution with 95% confidence interval (CI);
2. Estimate of model discrimination [Area under the receiver operating characteristic (ROC) Curve, AUC] with 95% CI of A) the non-*APOE* PRS alone; B) the PRS and *APOE* status; C) The non-genetic predictors alone; D) the full model.
3. Tail discrimination: We set the cutoff for the high risk group at the 97.5% of the PRS. The ORs and 95% CI (and the P-value for the OR) were calculated by comparing the high risk group vs. everybody else. i.e the participants in the top 2.5% of the PRS vs the bottom 97.5%.
4. The sensitivity / specificity as well as negative (NPV) and positive (PPV) predictive values at the proposed cutoff (split by ancestry if appropriate for each cohort).

The NPV/PPV used prevalence adjusted metrics, i.e. PPV = (Sn * Pr) / [(Sn * Pr) + ((1 – Sp) * (1 – Pr))] and NPV = (Sp * (1 – Pr)) / [(Sp * (1 – Pr)) + ((1 – Sn) * Pr)] where Sn = sensitivity, Sp = specificity, and Pr = population based prevalence reflective of the study population

### Proteomics study at two participant sites

The Brazilian Longitudinal Study of Adult Health (ELSA-Brasil) enrolled 15,105 civil servants aged 35 to 74 years living in six cities ^24^, addressing the incidence of non-communicable diseases. From the 15,105 participants, 9,333 DNA samples were analyzed for genetic ancestry using a software tool for maximum likelihood estimation of individual ancestries from multi-locus SNP genotype datasets^25^. The INTERVAL BioResource recruited 45,263 whole blood donors (22,466 men and 22,797 women) between June 11, 2012, and June 15, 2014^26^. Donors were aged 18 years or older from 25 NHS Blood and Transplant (NSHBT) blood donation centers distributed across England, UK. In addition to the GIRA validation at these two sites, the PRS scores were tested for association with 3,282 plasma protein targets from the SomaLogic proteomics platform (SomaLogic Inc., Boulder, CO, USA).

### Exploration of the PRS model in African populations

Without an African cohort of AD patients, we explored the distribution of the PRS scores in African populations in the Africa Wits-INDEPTH Partnership for genomic studies (AWI-Gen) leveraging its access via the IHCC resources^27^. African populations from three geographic regions (south - South Africa; east – Kenya; and west - Ghana/Burkina Faso) were assessed.

## Results

The genetic markers for the PRS model were polymorphic and informative in 4,145 ancestrally diverse dementia patients from the eMERGE consortium (Supplementary Table 2). Four cohorts from three sites, Korea Biobank, NCGG and East London Genes & Health, reported results of the PRS in dementia cases *vs*. controls (Table 2). Except the UKB white cohort, the best performance of prediction of dementia was by the full GIRA model with combination of the PRS component, the *APOE* haplotypes, and the non-genetic covariates. In the risk prediction, either genomic markers alone or non-genetic factors alone is insufficient, while the combined usage of genetic risk factors and non-genetic factors increased the performance of the prediction model. Most importantly, all four cohorts in Table 2 were non-European. A random effects restricted maximum likelihood (REML) meta-analysis of the four AUCs and their variance was 0.674 (0.643-0.706), indicating the potential of a prediction model in the non-European populations.

**Table 2.** Validation of PRS models (95% CIs)

| **Cohort** | **Odds ratio per SD** | **AUC with PRS only** | **AUC with genetic predictors (PRS and *APOE* haplotypes)** | **AUC with non-genetic covariates only** | **AUC with the full GIRA model\*** |
| --- | --- | --- | --- | --- | --- |
| **Korea pheno1** | 1.186<br>(0.992-1.418) | 0.548<br>(0.497-0.600) | 0.677(0.628-0.726) | 0.627(0.576-0.677) | 0.751(0.705-0.796) |
| **Korea pheno2** | 1.040(0.871-1.243) | 0.507(0.456-0.559) | 0.612(0.560-0.664) | 0.545(0.493-0.597) | 0.637(0.586-0.689) |
| **East London** | 1.110(0.940-1.330) | 0.530(0.470-0.590) | 0.540(0.480-0.600) | 0.680(0.610-0.750) | 0.690(0.620-0.760) |
| **Japan** | 1.120 | 0.545(0.520-0.570) | 0.607(0.582-0.632) | 0.616(0.591-0.641) | 0.625(0.601-0.650) |
| **UKB white** | 1.003(0.990-1.007) | 0.527(0.516-0.537) | 0.542(0.532-0.553) | 0.780(0.773-0.787) | 0.746(0.738-0.754) |

\*The full GIRA model: the model with PRS, APOE and non-genetic covariates.

Further, to assess the potential application of the PRS in African populations, the PRS model was examined in the continental African AWI-Gen cohort including participants from across the continent (https://www.wits.ac.za/research/sbimb/research/awi-gen/)^28^ ^29^ to determine if the SNP markers forming the PRS are polymorphic and informative in African populations from different geographic regions. As shown, the score is normally distributed with similar variance across African populations (Supplementary Figure 1), indicating it is suitable for use in populations of African ancestry.

In addition to the validation of the prediction model, two sites, ELSA Brazil and INTERVAL UK reported on results of the score as well as results from plasma proteomic analysis. The proteomics analysis showed correlation with *APOE* haplotypes, including also with blood APOE levels across two separate peptides (Table 3). Correlation of 99 proteins with the full PRS model were also observed with *P*<0.05 (Supplementary Table 3). Using the WebGestalt (WEB-based Gene SeT AnaLysis Toolkit) web tool^30^, over-representation analysis (ORA) of the correlated genes by the DisGeNET approach^31^, highlighted the genes (*CEBPB, FSHB, LEP, LHB, LIF*) in the gene set C0021361:Female infertility with false discovery rate (FDR)=0.0066779 (Table 4). ORA by the GLAD4U approach^32^ highlighted the gene sets PA445859:Thyroiditis, Autoimmune (FDR=0.017306, including *CD3G, CGA, CXCL9, FAS, LILRB1*); PA444172:Fetal Growth Retardation (FDR=0.044508, including *IGF1, IGFBP3, LEP, SELP, TMEM70*); PA443588:Cachexia (FDR=0.044508, including *IGF1, LEP, LIF, SELP*). Taken together, these results demonstrate enrichment of several genes that influence AD risk, as well as risks for multiple other conditions, including but not limited to female infertility, thyroiditis and autoimmunity. These results warrant further evaluation for risk optimization.

**Table 3.** Correlation of *APOE* haplotypes with APOE protein levels.

| **Proteomics ID** | **R2** | **P** | **BETA** | **SE** |
| --- | --- | --- | --- | --- |
| **APOE.2937.10.2**<br>**(isoform E3)** | 6.55E-03 | 0.0057 | 16.9 | 6.11 |
| **APOE.5312.49.3**<br>**(isoform E2)** | 0.00475 | 0.0194 | 15.03 | 6.42 |

**Table 4.** Genes in the gene-set C0021361:Female infertility associated with AD PRS.

| **Gene Symbol** | **PRS_R2** | **P** | **BETA** | **SE** | **Gene Name** |
| --- | --- | --- | --- | --- | --- |
| ***CEBPB*** | 0.004 | 0.032 | 15.645 | 7.274 | CCAAT enhancer binding protein beta |
| ***FSHB*** | 0.003 | 0.018 | -13.862 | 5.871 | follicle stimulating hormone subunit beta |
| ***LEP*** | 0.004 | 0.035 | -15.106 | 7.147 | leptin |
| ***LHB*** | 0.002 | 0.036 | -12.101 | 5.765 | luteinizing hormone beta polypeptide |
| ***LIF*** | 0.003 | 0.049 | -13.995 | 7.109 | LIF, interleukin 6 family cytokine |

## Discussion

As one of the first IHCC multinational studies, we demonstrate the feasibility of developing a trans-ethnic AD GIRA model that is predictive of disease predisposition across diverse populations, globally. In this regard, the IHCC project served as an important resource to examine GIRA and PRS in under-represented populations in genomic studies. The GIRA model performed better than the PRS model in East Asian populations from Japan and Korea, and in South Asian populations of Pakistani/Bangladeshi origin recruited through UK. While lacking a well phenotyped AD cohort of African origin with genetic data, we demonstrate a normal distribution of the PRS scores in different regions in Africa, suggesting that the current PRS score system is potentially informative in African populations. Further assessment of the PRS model in African AD patients is warranted.

In addition to its clinical potential, this study uncovered associations between the AD PRS scores and other disease-related gene sets, leading to novel insights into the pathogenesis of AD. In this regard, we identified association of the AD PRS and genes related to female infertility. Clinically, it has been observed that parity is inversely associated with risk of AD^33^. The genes related to female infertility identified in this study help explain the increased risk of AD in women, as well as the molecular mechanisms of the pathogenesis of AD in women and how parity decreases the risk of AD. Among the 5 genes related to female infertility identified (Table 4), the lowest P value was seen between PRS and lower level of follicle stimulating hormone subunit beta (encoded by *FSHB*). The physiological function of follicle-stimulating hormone is to induce egg and sperm production^34^. Genetic variants of *FSHB* have been shown to cause hypogonadotropic hypogonadism in women and men^35 36^. With The *FSHB* gene as a potential mediator of the pathogenesis of AD, it may also help to explain the clinical correlation between hypogonadism and AD^37^. More importantly, *FSHB* may represent a new opportunity to develop hormone therapy for AD^38 39^. In addition, previous studies have demonstrated correlations between abnormal levels of thyroid-stimulating hormone (TSH) and the risk of AD^40^. While further studies are warranted, our study identified 5 genes that may mediate this correlation.

Given that this is the first effort by the IHCC to leverage large existing datasets that reside within the IHCC consortium for a trans-ethnic GIRA on AD, we envision an opportunity to scale this to other cohorts within the consortium and expand the number of traits that can be analyzed. As such, the IHCC presents a rich resource of data for collaborative research with trans-ethnic focus, where there is much unmet need at present as this is an area of research that has been largely neglected. We also need to emphasize that this is a proof-of-principle study for an international effort to develop a trans-ethnic GIRA model for precision medicine, whereas to improve the GIRA model as suggested by the currently limited AUC scores is still warranted through further international efforts. As our research efforts continue, we envision efficient data sharing across academia and industry, where we will focus on improving patient health care services for AD and other diseases as well as biomedical research discoveries, leveraging our established GIRA approach.

## Data Availability

Supporting data from this study can be obtained by emailing the corresponding author Dr. Hakon Hakonarson.

## Supplementary materials

**Supplementary Table 1** SNPs and weights for the PRS model

**Supplementary Table 2** Frequencies of the genetic markers for the GIRA model in 4,145 ancestrally diverse dementia patients from the eMERGE consortium

**Supplementary Table 3** Correlation of 99 proteins with P<0.05 with the full GIRA model

**Supplementary Figure 1** Distribution of the PRS in the continental African AWI-GEN cohort from different geographic regions

## Acknowledgements

We thank all the participants who contributed to and enabled this study.

## Ethical Approval

This study was exempted by the Institutional Review Board (IRB) of the Children’s Hospital of Philadelphia, USA. Human participant personal information was not shared with the research group and participants were de-identification. All human participants or their proxies provided written informed consent for their respective studies and all the studies were approved by their local and/or national ethics review boards.

## Consent for publication

All authors have provided consent for publication of the manuscript.

