## Supplementary Figure 1 for "Trans-ethnic Genomic Informed Risk Assessment for Alzheimer’s disease: An International Hundred K+ Cohorts Consortium Study"

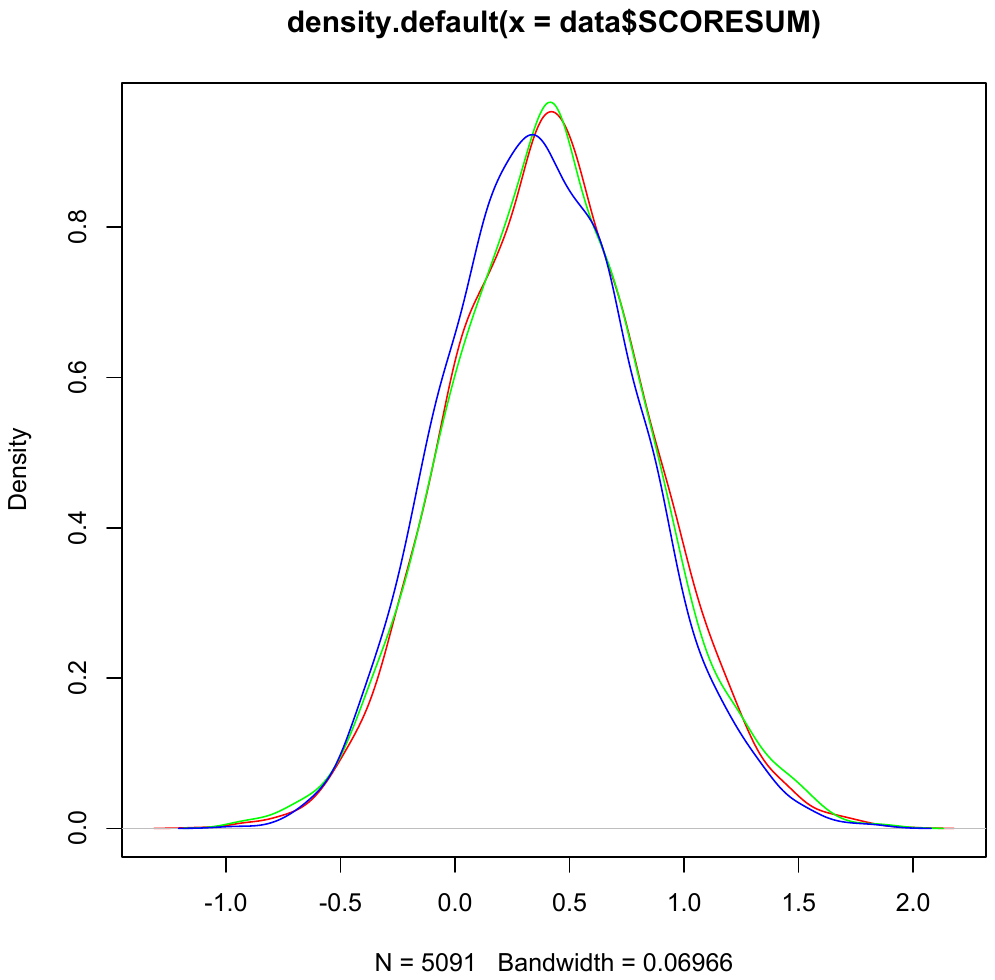

**Supplementary Figure 1. Distribution of the PRS in the continental African AWI-GEN cohort from different geographic regions.** Green - South Africa; red – Kenya; and blue - Ghana/Burkina Faso.
